# Resting energy expenditure of women with and without polyendocrine metabolic ovarian syndrome: a systematic review and meta-analysis

**DOI:** 10.64898/2025.12.03.25341536

**Authors:** Richard Kirwan, Leigh Peele, Gregory Nuckols, Georgia Kohlhoff, Hannah Cabré, Alyssa Olenick, James Steele

## Abstract

Context: Polyendocrine metabolic ovarian syndrome (PMOS; previously polycystic ovarian syndrome i.e., PCOS) is common in reproductive-age women, who often have higher BMI classification. This is assumed to stem from lower resting energy expenditure (REE), influencing lifestyle intervention guidelines. However, evidence for reduced REE in women with PMOS compared with those without is inconsistent. Objective: To systematically search and meta-analyse the existing literature to estimate and describe the difference in REE between women with and without PMOS. Data Sources: PubMed, MEDLINE/EBSCO, and Web of Science were searched for published research from January 1990 through May 2025, and the searches were updated through September 2026. The update identified no additional eligible studies. Study Selection: Studies that measured REE in women living with PMOS, both with and without control arms of women without PMOS, were included. Data Extraction: Bibliometric, demographic, and REE data were extracted by one investigator and checked in triplicate. Data Synthesis: Thirteen studies were included in a Bayesian arm-based multiple condition comparison (i.e., network) type meta-analysis model with informative priors to compare both mean REE, and between-person variation in REE, between women with and without PMOS. Mean REE differed between groups by 31 kcal/day [95% quantile interval: −44 to 113 kcal/day] and the contrast ratio for between-person standard deviations was 0.98 [95% quantile interval: 0.71 to 1.32]. Conclusions: These findings indicate that REE does not meaningfully differ between women with and without PMOS. Group-level differences in resting energy expenditure are small, insignificant, or not physiologically relevant.

## Introduction

Polyendocrine metabolic ovarian syndrome (PMOS^1^) affects approximately 10% of women of reproductive age worldwide, making it the most common endocrine disorder affecting this population^2^. Due to several factors including hyperandrogenism and alterations in insulin resistance, PMOS is believed to contribute to an increased risk of diabetes, metabolic syndrome and cardiovascular disease^3–7^, along with being a leading cause of anovulatory infertility in women^8^. Furthermore, epidemiological data has consistently demonstrated that women with PMOS are significantly more likely to suffer from overweight or obesity, compared to the general female population, with estimates ranging from 38% to 88% of PMOS patients falling into overweight or obese body mass index (BMI) categories^9,10^.

The elevated incidence of overweight and obesity in PMOS is likely multifactorial with proposed mechanisms including blunted postprandial appetite hormone responses leading to reduced satiety and increased food cravings^11–13^ and a reduced resting energy expenditure (REE)^14^. Indeed, the study from Georgopoulos et al.^14^ examined REE in women with and without PMOS using indirect calorimetry and reported that resting energy expenditure was approximately ~400 kcal/day lower in women with PMOS. Notably, they also reported that insulin resistance further reduced REE among women with PMOS, with insulin-resistant women exhibiting an additional reduction nearly 500 kcal per day compared to women with PMOS who were not insulin resistant^14^. Other studies similarly report lower REE in women with PMOS using indirect methods (such as prediction from bioelectrical impedance analysis or accelerometer physical activity data), also suggesting that factors including insulin resistance and BMI category influence REE in women with PMOS^15,16^. However, despite the widespread acceptance that women living with PMOS exhibit reduced REE based on studies such as these, other research has reported little to no difference in REE between women with and without PMOS^17–19^.

The consequences of widespread acceptance that REE is substantially lower in women living with PMOS should not be underestimated, particularly in light of the aforementioned incidence of overweight and obesity in this population. Women with PMOS typically engage in more frequent weight-loss attempts than women without PMOS^20^. From a physiological perspective, if women with PMOS do exhibit a lower REE, this could imply a meaningful metabolic disadvantage that may influence dietary and nutritional guidance for weight management; for example, recommending a slightly more severe energy restriction to overcome the belief that they have a lower REE^21^. Recommendations such as this could influence the well documented prevalence of eating disorders in women with PMOS^22,23^. Contrastingly, belief in a “slower metabolism” could instead serve as a deterrent to energy restriction based weight-loss approaches for some women in line with typical general population recommendations that are also recognised as efficacious for improving PMOS symptoms^24,25^ and are routinely recommended^26,27^. It has been well documented that women with PMOS already experience higher rates of anxiety, depression, and lower quality of life (QOL) as a result of negative body image and weight-related concerns^28–31^. Clarifying the relationship between REE and PMOS may therefore help guide more accurate clinical recommendations and empower both practitioners and women with PMOS.

A recent scoping review has highlighted the inconsistency in research examining energy expenditure in women with PMOS^32^. However, to our knowledge there have been no attempts to systematically review, and quantitatively synthesise via meta-analysis, studies of REE in women with, and without, PMOS. Therefore, to estimate and describe the magnitude of difference in REE between women with and without PMOS, we completed a systematic review and meta-analysis of studies reporting REE in these populations. Our primary aim was to estimate the difference in mean REE between women with and without PMOS. A secondary aim was to compare between-person variability in REE between the groups.

## Materials and Methods

This systematic review and meta-analysis was pre-registered on PROSPERO (CRD42024601434) initially on the 3^*rd*^ of December 2024 and performed in accordance with the Preferred Reporting Items for Systematic Reviews and Meta-Analyses (PRISMA) statement guidelines^33^. The updated PRISMA flow diagram reported below (Figure 1) was generated using a reproducible R script in the accompanying repository. The primary aim of this review was to examine the descriptive question “Does resting energy expenditure (REE) differ between women with and without polyendocrine metabolic ovarian syndrome (PMOS)?”. We summarise and describe the studies in addition to quantitatively synthesising their results via meta-analysis.

**Figure 1:**
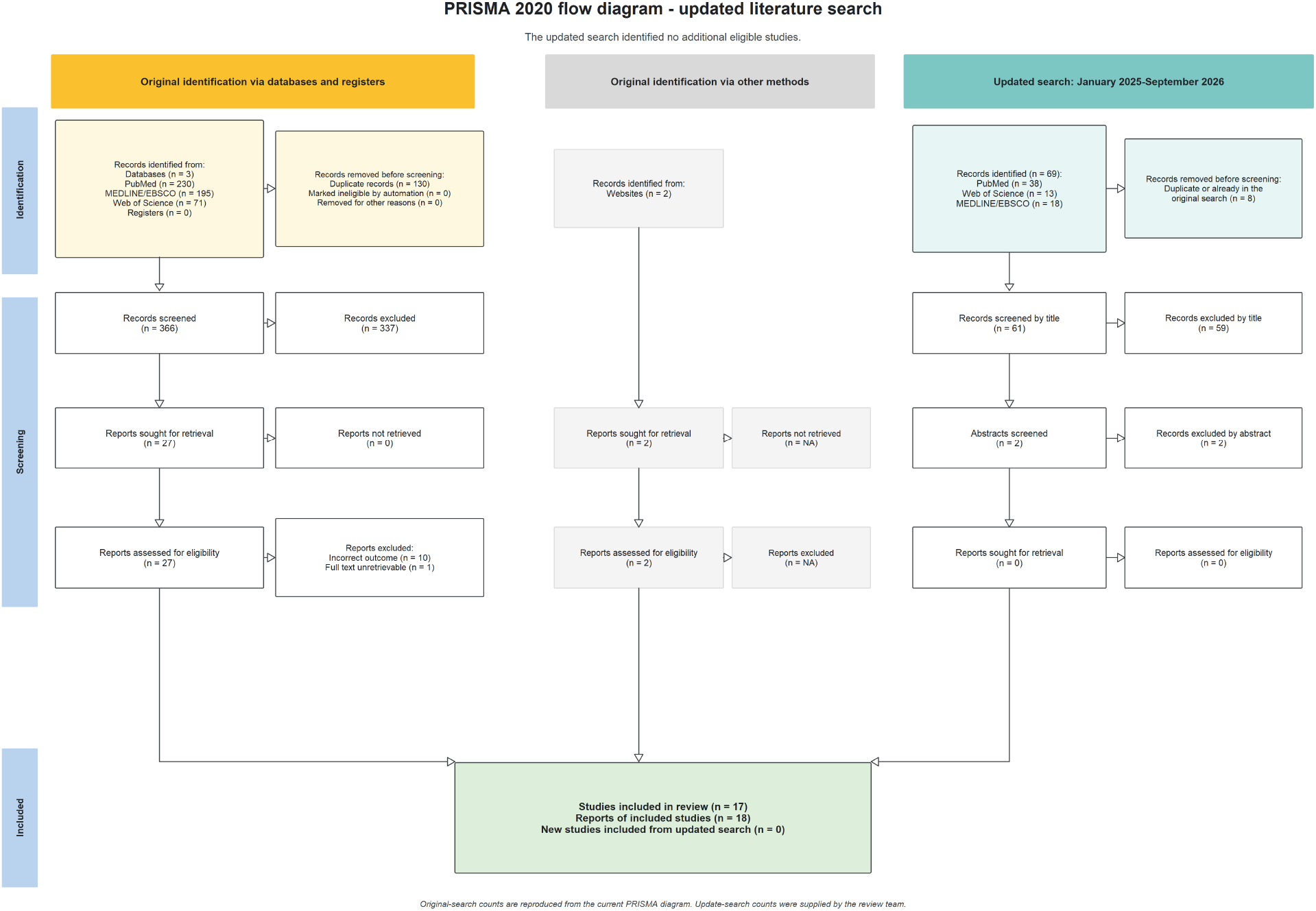
Updated PRISMA 2020 flow diagram for the systematic review. The updated database search covered January 2025 through September 2026 and identified no additional eligible studies. The website shown in the original-search pathway was a prior narrative review by members of the author group that identified two studies not found in the systematic database search.

### Search Strategy

PubMed, Web of Science, and MEDLINE/EBSCO were searched using the following Boolean search string: ((“Basal Metabolic Rate”[MeSH] OR “Energy Metabolism”[MeSH] OR “Resting Metabolic Rate” OR RMR OR “Resting Energy Expenditure” OR REE OR “Basal Metabolic Rate” OR BMR OR “resting energy” OR “basal energy expenditure”) AND (“Polycystic Ovary Syndrome”[MeSH] OR “Polycystic Ovary Syndrome” OR PCOS OR “Polycystic Ovarian Disease” OR “Stein-Leventhal Syndrome”)). The initial search covered publications from January 1990 through May 2025. The searches were subsequently updated for the deliberately overlapping period January 2025 through September 2026 to ensure continuity at the boundary of the original search. The global consensus adopting “Polyendocrine Metabolic Ovarian Syndrome” (PMOS) was published only in June 2026, during the final months of the update^1^. For continuity with the original strategy and the terminology under which the historical literature was indexed, the updated strategy retained the reported PCOS terminology and did not add PMOS. The update identified 69 records: 38 in PubMed, 13 in Web of Science, and 18 in MEDLINE/EBSCO. Eight duplicate records, including records already captured by the original search, were removed, leaving 61 records for title screening. Fifty-nine were excluded at title screening and the remaining two were excluded after abstract screening. No reports proceeded to full-text retrieval and no additional eligible studies were identified; consequently, the included-study set and all analyses were unchanged. Searches were limited to English-language articles, and Rayyan was used to manage the search and screening process. Two reviewers (RK and GK) independently screened all titles and abstracts against the predefined inclusion and exclusion criteria. Articles deemed potentially eligible by either reviewer were retrieved in full text. Full texts were then independently assessed by RK and LP to determine final eligibility. Any disagreements at either stage were resolved through discussion, and when consensus could not be reached, a third reviewer acted as an adjudicator.

### Eligibility Criteria

Studies were included in the systematic review if 1) participants were confirmed as women with PMOS between the ages of 18 to 65 years of age with or without insulin resistance; 2) participants were otherwise healthy (e.g., non-diabetic, no cardiovascular disease); 3) studies reported REE measured using any valid method, such as direct or indirect calorimetry or doubly labelled water; and 4) trials were not retracted at the time of this analysis. Studies were excluded if they 1) used invalid or non-standard methods for measuring REE (e.g., predicted REE from body composition or accelerometer data); 2) were non-peer-reviewed journal articles (including grey literature sources such as conference abstracts, theses and dissertations); or 3) were secondary analyses with the same primary outcome data as another included study.

The condition being studied was PMOS and we included observational cross-sectional design studies, in addition to intervention studies where REE was reported for the population (and if present, the comparator i.e., women without PMOS) condition of interest. For clarity, studies of any design were included if they reported the REE using the methods indicated for a sample of adult women with PMOS and who were otherwise healthy. This included both studies with and without samples of healthy control women without PMOS. As detailed in the statistical analysis section below, a Bayesian model with informative priors based on normative data for REE in healthy women without PMOS was included to provide control information indirectly where this was missing. The use of such priors is an efficient tool for incorporating historical information on a particular population in a conservative manner^34^.

Following the PICO framework our eligibility criteria can be defined as follows:

- Population
  - Inclusion criteria:
    * Women
    * 18-65 y
    * With or without insulin resistance (IR)
- Intervention(s) or exposure(s)
  - Otherwise healthy women with PMOS
- Comparator(s) or control(s)
  - Otherwise healthy control women without PMOS
- Outcome
  - Inclusion criteria
    * Resting energy expenditure (REE) measured using any valid method including direct/indirect calorimetry, doubly labelled water.
  Exclusion criteria:
    * Studies using invalid or non-standard methods for measuring REE (e.g., predicted REE from body composition or accelerometer data)

### Data extraction (selection and coding)

Data were extracted by one investigator and checked in triplicate (i.e., were separately checked against the included studies three times by the investigator). No formal piloting of data extraction was performed but, given the number of identified studies was not overwhelmingly large, the process of triplicate extraction was used to develop and refine the Excel spreadsheet used for data extraction. Where there were any ambiguities or missing data that were required for our analysis, corresponding authors of the included articles were contacted to clarify or provide this information.

Bibliometric data including authors, journal, and article titles were extracted. Descriptive statistics for age, body mass, fat mass, fat-free mass, height, BMI, race, physical activity levels, country of investigation, information regarding glucose/insulin regulation and insulin resistance status (where available), diagnostic criteria for PMOS, and measurement method and device were extracted for each arm within each study in addition to sample size. Descriptive characteristics were then tabulated across studies for reporting. For each arm, and observation time point if multiple observations reported (e.g., before and after an intervention), depending on what was reported by the authors we extracted the means, medians, standard deviations, standard errors, lower and upper range values, and interquartile range for the unadjusted and/or body-mass-adjusted and/or fat-free-mass-adjusted REE values. Where REE values adjusted for body mass and/or fat-free mass were reported we used the reported body mass and/or fat-free mass mean values for that arm to convert them to unadjusted REE values (i.e., multiplied them by body mass and/or fat-free mass mean values). Where means and/or standard deviations were missing the latter were either calculated from standard errors and sample size, or both were estimated from lower and upper range, interquartile range, median, and sample size depending on the available information using the methods of Wan et al.^35^. Further, where missing, height, body mass, or BMI were estimated based on the reported means. All extracted REE values were converted to kcal/day. In only one case^36^ REE was reported relative to body mass and the unadjusted values were no longer available (confirmed by the authors). In this case, we used the mean body mass to estimate unadjusted REE.

### Studies with possible reporting errors

During data extraction it was noted that several studies from the same lab/research group^14,37–39^ contained a number of discrepancies that seemed to be possible reporting errors. These included, based on taking the authors’ results as written, standard errors that implied impossible or at least incredibly unlikely standard deviations, and discrepancies in sample size reporting throughout for most variables without explanation or where this was explained the sample sizes were discrepant with the text. Further, data were not reported for the healthy control women without PMOS in three of the studies^37–39^, and REE was reported as an “adjusted” value whereby REE_*adjusted*_ = REE_*group mean*_ +(REE_*adjusted*_ ™REE_*predicted*_) and the REE_*predicted*_ was obtained by substituting the individual lean body mass, fat mass, gender, and age in the linear regression equation generated by the data of all patients. In correspondence with the senior author we were unable to clarify the reporting discrepancies as the person responsible for the data/results was no longer contactable. The original data were also no longer available, so we could not calculate the unadjusted REE.

Given these issues we decided to extract the results from these studies as reported and to conduct analyses both with and without their inclusion. Though not pre-registered, due to a lack of confidence in the reported results, we decided to include the analysis omitting these studies as our main models in the results reported below. The results of the analysis including them are reported in the sensitivity analysis section.

### Statistical Analysis

#### Analytical overview

The primary analysis estimated the difference in mean resting energy expenditure (REE), expressed in kcal/day, between women with PMOS and women without PMOS. Positive values indicate higher mean REE in the PMOS group. We used the pre-registered Bayesian arm-based meta-analysis because some eligible studies included both groups, whereas others included only women with PMOS, and some studies contributed more than one REE measurement. Unlike a conventional pairwise meta-analysis, which can use only studies containing both groups, the arm-based approach allowed all eligible study arms to contribute information while accounting for the dependence between multiple estimates from the same study or research group.

Information from large studies of REE in women without PMOS was used to define plausible values for the control population^34,40^. The prior distribution for the difference between groups was centred on no difference and allowed a wide range of values in either direction. The difference in mean REE was the primary estimand. As a secondary analysis, we compared between-person variability in REE using the ratio of the standard deviations in the PMOS and control groups; a ratio of 1 indicates similar variability.

We summarised posterior distributions using their means and equal-tailed 95% quantile intervals (QI). The pre-registered sensitivity analysis was a pairwise meta-analysis restricted to studies that directly compared women with PMOS and controls at baseline. This analysis used weakly regularising priors and did not incorporate single-arm evidence or historical control information. Additional sensitivity analyses examined the effect of including four studies with unresolved reporting concerns and of restricting intervention studies to baseline measurements. Exploratory models examined adjustment for study-level mean BMI or fat-free mass. The complete model equations, prior distributions, deviations from the pre-registration, computational checks, post-processing procedures, and full results are reported in the Supplementary Technical Analysis and Results. All analysis code is available in the accompanying repository at https://github.com/jamessteeleii/pcos_ree_meta.

#### Use of generative artificial intelligence

OpenAI ChatGPT/Codex (GPT-5.6 Sol; accessed September 2026) was used as an author-directed computational assistant during a post-submission audit of the analysis code and reproducible workflow and in the development of the supplementary bibliometric analysis. The tool assisted with code review and development, identification of potential implementation errors that contradicted the pre-registered analysis plan, construction of automated validation checks, workflow documentation, and the organisation and classification of publicly available bibliometric and web-source information. All AI-assisted code and outputs were reviewed, revised, executed, and validated by JS against the preregistration, canonical source data, original publications and webpages, automated reproducibility checks, and model diagnostics. Final decisions concerning the data, methods, model specifications, analyses, source inclusion and classification, and interpretation were made by the authors. The computational audit resulted in minor changes to some reported numerical estimates but did not alter the review question, principal estimands, substantive interpretations, or conclusions. The bibliometric work informed only the ancillary descriptive account of the dissemination of included studies. No confidential or participant-level data were provided to the AI system.

## Results

### Included studies

The systematic review identified 17 eligible studies. The updated search through September 2026 identified no additional eligible studies, so the review and analysis populations were unchanged. Four studies contained unresolved reporting concerns and were retained in the descriptive review but omitted from the primary analysis; they were reinstated in a sensitivity analysis. The primary analysis therefore included 13 studies, 24 arms (control: 9; PMOS: 15), and 918 participants at baseline. The flow from study identification to each synthesis is shown in Figure 1, and complete study characteristics are provided in the online descriptive table.

All but one included study measured REE using indirect calorimetry; one used doubly labelled water. The women with PMOS represented in the available evidence were generally young, had a higher BMI than controls, and the groups with sufficient metabolic information were considered insulin resistant. The applicability of the findings to lean or insulin-sensitive PMOS phenotypes is therefore uncertain.

### Primary and secondary analyses

In the primary arm-based analysis, mean REE was estimated to be 31 kcal/day higher in women with PMOS (95% QI: −44 to 113 kcal/day). The evidence was therefore compatible with small differences in either direction but did not support a substantial reduction in REE among women with PMOS. In the secondary analysis, between-person variability was also similar between groups: the PMOS-to-control ratio of standard deviations was 0.98 (95% QI: 0.71 to 1.32).

The posterior distributions and study-level conditional estimates for mean REE are presented in Figure 2, and the corresponding results for the between-person standard deviation of REE are presented in Figure 3.

**Figure 2:**
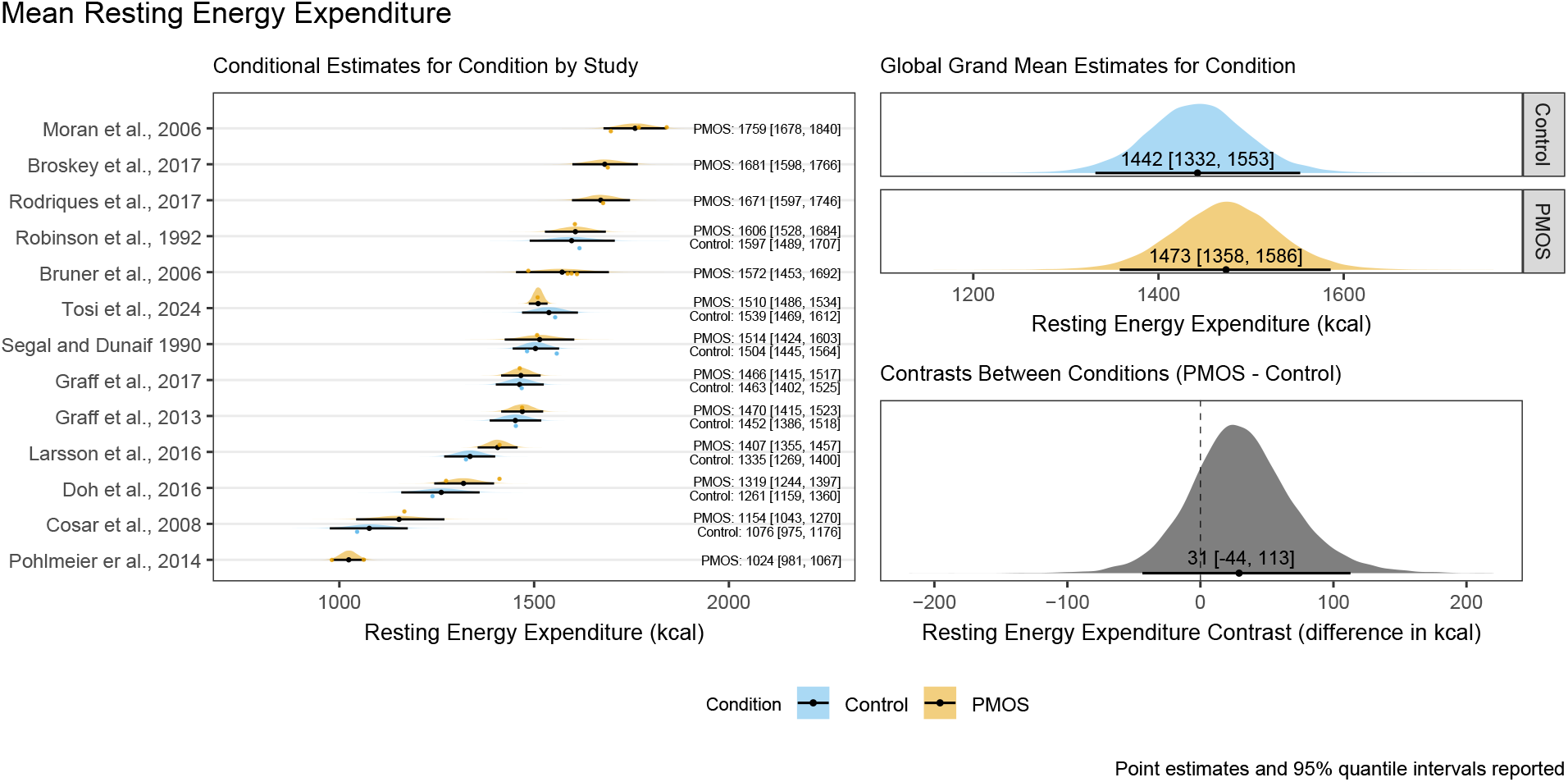
Posterior distributions, point estimates, and 95% quantile intervals for the study-level conditional estimates, global estimates by condition, and the contrast between conditions for mean resting energy expenditure.

**Figure 3:**
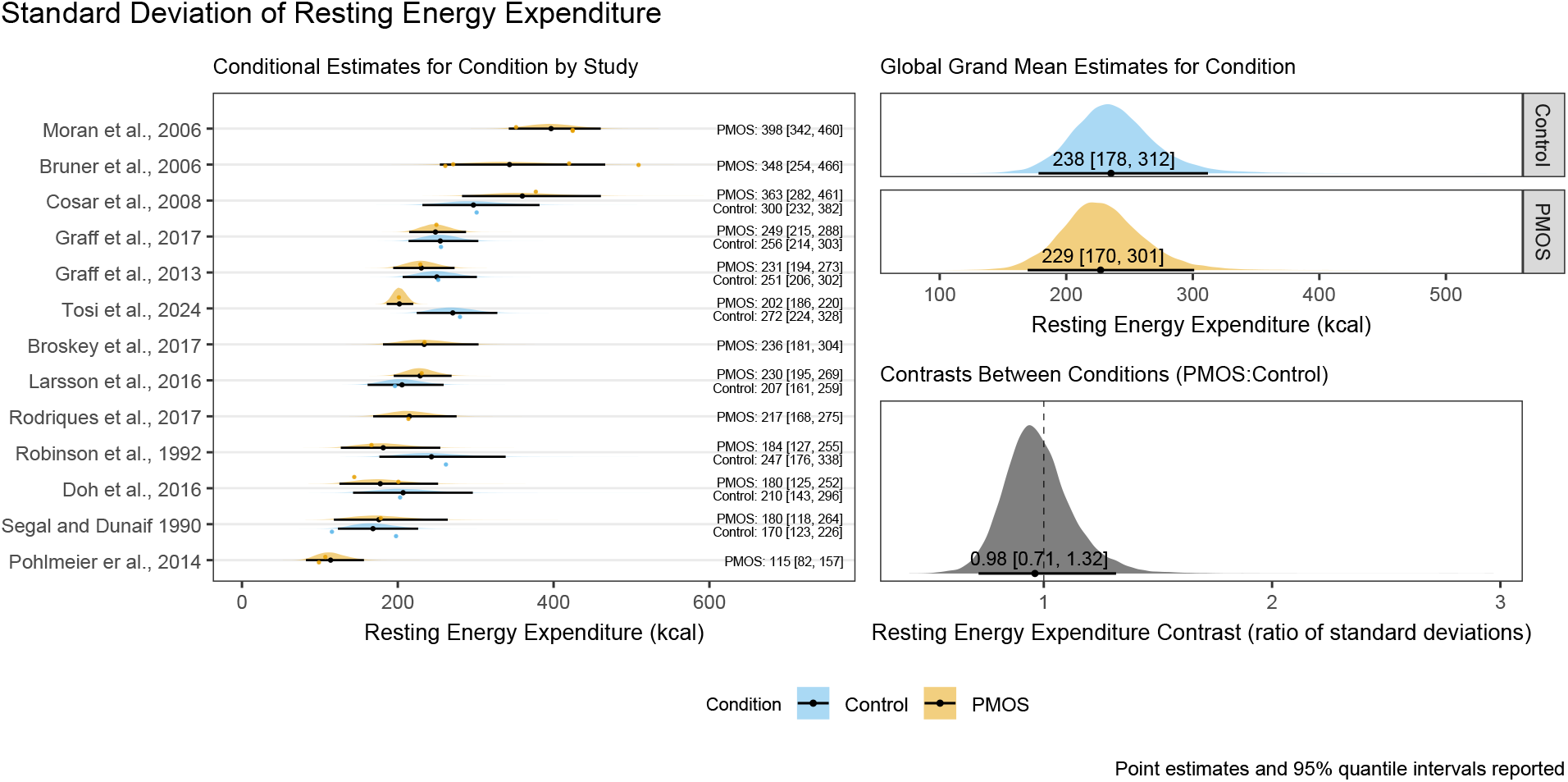
Posterior distributions, point estimates, and 95% quantile intervals for the study-level conditional estimates, global estimates by condition, and the contrast ratio between conditions for the between-person standard deviation of resting energy expenditure.

### Sensitivity and exploratory analyses

**Table 1:**
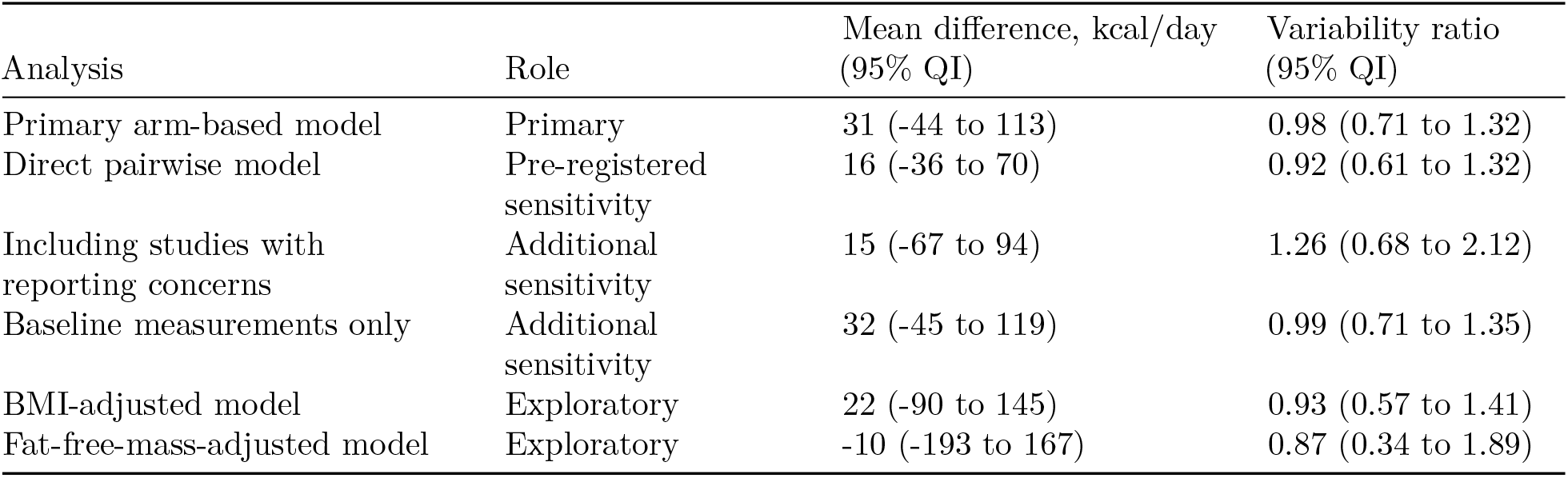
Summary of primary, sensitivity, and exploratory analyses.

The pre-registered pairwise analysis, restricted to studies that directly compared women with PMOS and controls at baseline, estimated a mean difference of 16 kcal/day (95% QI: −36 to 70). This result gives greater prominence to the direct comparisons and shows that the overall interpretation did not depend on incorporating single-arm studies or historical control information. None of the additional sensitivity or exploratory analyses materially altered the conclusion. In the table, variability is expressed as a PMOS-to-control standard-deviation ratio except for the direct pairwise model, which uses a coefficient-of-variation ratio. Complete estimates, conditional predictions, posterior visualisations, and technical interpretation are provided in the [Supplementary Technical Analysis and Results](https://jamessteeleii.github.io/pcos_ree_meta/supplementary_technical_analysis_results.html).

## Discussion

This study sought to estimate and describe the magnitude of difference in REE between women with and without PMOS. Most studies identified in the systematic review, and included in the meta-analysis, used indirect calorimetry as the primary measure of REE and assessed women with PMOS who were insulin resistant and categorised as being in overweight or obese BMI categories compared to healthy controls. Our results indicate there is only a small magnitude of difference in REE (31 kcal/day [95% QI: −44 kcal/day to 113 kcal/day]) between women with PMOS and those without. Further, there is little difference in between-person variation between the groups based on the ratio of standard deviations (0.98 [95% QI: 0.71 to 1.32]) suggesting that, despite individual differences in REE, PMOS is not systematically associated with lesser or greater individual variability.

These findings challenge the widely held belief^2^ that PMOS is inherently associated with a slower metabolism^27^, predisposing women with PMOS to weight gain. The mistaken belief that REE is lower for those with PMOS may have led to recommendations centred on slightly more severe calorie restriction to achieve weight-loss goals, compared with recommendations for the general population, as primary management strategies for women with PMOS^21^. Recognising that there may be minimal differences in REE between women with PMOS and those without can inform both clinical and public practices, potentially leading to a shift in focus away from a requirement for more severe caloric restriction as a primary method of treatment towards more comprehensive, individualised, and psychologically safe approaches to care^26,27^.

The available evidence does not support substantially lower REE as an explanation for weight-related differences observed in women with PMOS given small group-level differences between women with PMOS and those without (1473 kcal/day [95% QI: 1358 kcal/day to 1586 kcal/day] versus 1442 kcal/day [95% QI: 1332 kcal/day to 1553 kcal/day], respectively). If anything, REE may be slightly higher in women with PMOS compared to healthy women without PMOS. BMI in women with PMOS ranged from 26.4 to 39.9 and was typically greater than in women without PMOS, in whom it ranged from 20.5 to 32.3; this may explain the slightly greater REE in the former group. However, for those studies where we could extract or estimate BMI, our additional exploratory models adjusted for this similarly showed little difference in REE (22 kcal/day [95% QI: −90 kcal/day to 145 kcal/day]) between women with PMOS and those without. However, one of the studies included in our analysis^41^ reported that, whilst there was little difference in unadjusted REE, when REE was adjusted for fat-free mass it was lower in women with PMOS, suggesting the potential importance of fat-free tissue in energy regulation. Yet, for those studies where fat-free mass was reported, our additional exploratory models adjusted for this similarly showed little difference in REE (−10 kcal/day [95% QI: −193 kcal/day to 167 kcal/day]) between women with PMOS and those without. As such, even adjusted for both BMI and fat-free mass, there seems to be little difference in REE between women with and without PMOS. Yet, a recent systematic review of mechanisms for metabolic dysfunction has reported that excess androgen may drive metabolic issues within adipose tissue and muscle tissue, contributing to complications such as obesity and insulin resistance^42^. Taken together, these findings highlight that factors beyond REE and typical correlates of this including BMI or fat-free mass, such as hormonal and tissue-specific metabolic effects, may play a more significant role in weight regulation challenges in women with PMOS.

As noted, women with PMOS are more likely to engage in weight-loss attempts^20^ and there could be concerns that this could inadvertently further foster the already well documented disordered eating in this population^22,23,43,44^. The pathways linking PMOS and disordered eating are multifactorial. Biological mechanisms such as hyperandrogenism, hyperinsulinaemia, and altered ghrelin and leptin signalling can heighten hunger, carbohydrate cravings, and appetite variability^45^. Frequent hypoglycaemia and associated mood changes have also been observed, which can trigger compensatory eating or binge episodes^45^. These physiological processes interact with psychological and social stressors, including infertility concerns, conflicting nutrition advice, chronic dieting, and exposure to idealised body images on social media, which together compound vulnerability to disordered eating^46^. Moreover, eating disorders themselves can disrupt endocrine function, potentially worsening PMOS symptoms and creating a self-reinforcing cycle^44,47^.

Understanding the intertwined biological, psychological, and social influences suggests the importance of considering whether restrictive dietary advice is appropriate given it may exacerbate feelings of failure, hunger dysregulation, and shame^48^. These concerns, coupled with the lack of difference in REE between women with and without PMOS might suggest that energy restriction based dietary interventions for weight-loss may be unnecessary. But, there is also evidence supporting the effect of energy restricted dietary interventions for improving PMOS symptoms^24,25^ and they are recommended in international guidelines^26,27^. Encouragingly though, these guidelines also recognise weight stigma as a determinant of health and call for its reduction across clinical and public health settings. Evidently the greater prevalence of women with PMOS falling into overweight and obese BMI categories compared to women without PMOS^9,10^ is unlikely to be due to differences in REE and so, despite the potential effectiveness of energy restriction dietary interventions, there is potential value in moving towards more weight-neutral, individualised, and empowering care following holistic guidelines recommending multiple approaches to management^21,26,27^.

A weight-neutral dietary approach emphasises promoting cardiometabolic health by focusing on adding or emphasising healthful foods instead of restricting intake or targeting weight loss explicitly. Instead of pursuing energy deficits, setting specific energy intake goals, eliminating food groups such as dairy, gluten, or carbohydrates, or labelling foods as ‘good’ or ‘bad’ for PMOS or weight loss, a clinician or dietitian might guide individuals to plan meals rich in protein, fibre, fruits, vegetables, legumes, and unsaturated fats, aligned with Mediterranean or DASH diet principles. This strategy should be tailored to individual preferences and avoid unnecessarily eliminating foods, which could reinforce restrictive behaviours, especially in populations at risk for disordered eating. Monitoring might include blood lipids, cholesterol, glucose, insulin, blood pressure, or menstrual cycle regularity, rather than focusing solely on body weight^27,49^.

### Strengths and Limitations

The current study has multiple strengths stemming from its preregistered, comprehensive methodology and Bayesian statistical approach. This statistical framework allowed us to incorporate studies with and without control groups to better estimate REE in women with and without PMOS and to perform multiple sensitivity analyses that confirmed the stability of our findings. However, a limitation here is that variability in methods across studies, such as differences in PMOS diagnostic criteria and REE testing protocols, may have influenced results and the relatively small number of studies overall limits the extent to which we can explore these potential moderators. Furthermore, some studies controlled for body weight or body composition when reporting REE values, while others did not. We accounted for this by estimating or converting reported data to obtain unadjusted REE values across all groups, thereby reducing this variability and further as noted above provided estimates adjusted for BMI and fat-free mass both of which had little influence on our conclusions. Another limitation is that, due to fewer total control groups than PMOS groups, informed priors were required in several statistical models. However, in the context of Bayesian meta-analysis this can also be considered a strength. Furthermore, our pairwise sensitivity model used weakly regularising default priors and produced a similar estimate, suggesting that the conclusions were not driven by the historical priors. Additionally, some studies reported data inconsistencies that could not be clarified (e.g.,^14,37–39^) and were dropped from our main analysis though our conclusions again did not qualitatively change when we conducted sensitivity analyses including these studies. Most included studies were cross-sectional or baseline assessments within intervention trials, which limits causal inference. Indeed, we did not pre-register any kind of causal model (e.g., a directed acyclic graph) to inform our analysis approach for causal inference and as such have been explicit about the estimates presented as being descriptive. Additionally, a formal risk-of-bias assessment was not undertaken. The available studies contributed heterogeneous forms of descriptive evidence, including cross-sectional comparisons, single-arm observations and repeated measurements from intervention studies, whereas commonly used risk-of-bias tools principally assess causal intervention or exposure effects. We therefore addressed limitations directly relevant to the descriptive REE estimates through detailed examination of diagnostic and measurement methods, explicit documentation of reporting inconsistencies and sensitivity analyses excluding studies with unresolved data concerns and post-baseline intervention measurements. These procedures are not equivalent to a formal risk-of-bias assessment, however, and residual bias arising from participant selection, measurement, missing information or selective reporting cannot be excluded. Our search was also restricted to PubMed, MEDLINE/EBSCO, and Web of Science; the overlap between PubMed and MEDLINE and the absence of databases such as Embase or CINAHL may have limited coverage, although the updated search through September 2026 identified no additional eligible studies. Finally, the women with PMOS included in the available studies were generally young, had higher BMI than control participants, and the groups with sufficient metabolic information were considered insulin resistant. The findings may therefore not necessarily apply equally to lean or insulin-sensitive PMOS phenotypes.

## Conclusion

In conclusion, the findings from this meta-analysis indicate that REE does not meaningfully differ between women with and without PMOS. Group-level differences in REE were small, insignificant, or not physiologically relevant. Additionally, variability in REE between individuals was also similar. These results suggest that a lower baseline REE is not associated with the weight-related challenges often associated with PMOS. These findings challenge the popular narrative that women with PMOS have a lower REE and may help better inform dietary interventions and nutritional support for these individuals. Future research should include more standardized REE measurement and reporting protocols, greater data transparency, consistent control and reporting of body weight or body composition, the presentation of both absolute and relative REE, and more precise characterization of PMOS phenotypes. Overall, these findings support the conclusion that PMOS is not negatively associated with REE and may help practitioners and researchers focus on individually targeted and holistic lifestyle interventions rather than negatively framed interventions based on unsupported assumptions regarding REE.

## Acknowledgements

The authors acknowledge the use of OpenAI ChatGPT/Codex (GPT-5.6 Sol; accessed September 2026) to assist with substantial editing and organisation of the revised manuscript, response to reviewers, technical documentation, and supplementary materials. The tool was used to generate the initial reviewer response document structure and to help audit consistency between the manuscript, analytical code, tables, figures, and reported results. All AI-assisted suggestions were critically reviewed, revised, and approved by the authors, who retained intellectual control over the work and take full responsibility for its accuracy and integrity.

## Data Availability

All data and code utilised for data preparation, transformations, analyses, plotting, and reporting are available in the corresponding GitHub repository https://github.com/jamessteeleii/pcos_ree_meta.

## Contributions

Gregory Nuckols and Leigh Peele conceived the idea for the project. All authors contributed to the design of the project and methods. Richie Kirwan and Leigh Peele conducted the systematic search and screening. James Steele performed the data extraction, conducted the statistical analyses, and produced the data visualisations. All authors contributed to drafting the initial manuscript. All authors contributed to editing the manuscript. All authors read and approved the final manuscript.

## Footnotes

1 Recent consensus guidelines have renamed what was previously called polycystic ovarian syndrome (PCOS) to Polyendocrine metabolic ovarian syndrome (PMOS)^1^. We adopt the new terminology throughout the manuscript though note that this project was initiated and searches conducted prior to this change being adopted.

2 We conducted a [supplementary bibliometric analysis](https://jamessteeleii.github.io/pcos_ree_meta/supplementary_bibliometric_impact.html) with ChatGPT/Codex (GPT-5.6 Sol version) to explore the scholarly, extended scholarly, grey-literature, and public dissemination and discourse of the studies included in our review in order to understand descriptively how prominent certain articles and findings have been. The findings of this supplementary analysis do indicate that, whilst not the most highly cited in scholarly works, the 2009 study from Georgopoulos et al. that reported a significantly reduced BMR in women with PMOS^14^ has been widely cited and reinforced in both academic and particularly public contexts.

